# Study Protocol for the Artificial Intelligence-Driven Evaluation of Structural Heart Diseases Using Wearable Electrocardiogram (ID-SHD)

**DOI:** 10.1101/2024.03.18.24304477

**Authors:** Arya Aminorroaya, Lovedeep Singh Dhingra, Aline Pedroso Camargos, Sumukh Vasisht Shankar, Akshay Khunte, Veer Sangha, Sounok Sen, Robert L McNamara, Norrisa Haynes, Evangelos K Oikonomou, Rohan Khera

## Abstract

**Introduction:** Portable devices capable of electrocardiogram (ECG) acquisition have the potential to enhance structural heart disease (SHD) management by enabling early detection through artificial intelligence-ECG (AI-ECG) algorithms. However, the performance of these AI algorithms for identifying SHD in a real-world screening setting is unknown. To address this gap, we aim to evaluate the validity of our wearable-adapted AI algorithm, which has been previously developed and validated for detecting SHD from single-lead portable ECGs in patients undergoing routine echocardiograms in the Yale New Haven Hospital (YNHH).

**Research Methods and Analysis:** This is the protocol for a cross-sectional study in the echocardiographic laboratories of YNHH. The study will enroll 585 patients referred for outpatient transthoracic echocardiogram (TTE) as part of their routine clinical care. Patients expressing interest in participating in the study will undergo a screening interview, followed by enrollment upon meeting eligibility criteria and providing informed consent. During their routine visit, patients will undergo a 1-lead ECG with two devices - one with an Apple Watch and the second with another portable 1-lead ECG device. With participant consent, these 1-lead ECG data will be linked to participant demographic and clinical data recorded in the YNHH electronic health records (EHR). The study will assess the performance of the AI-ECG algorithm in identifying SHD, including left ventricular systolic dysfunction (LVSD), valvular disease and severe left ventricular hypertrophy (LVH), by comparing the algorithm’s results with data obtained from TTE, which is the established gold standard for diagnosing SHD.

**Ethics and Dissemination:** All patient EHR data required for assessing eligibility and conducting the AI-ECG will be accessed through secure servers approved for protected health information. Data will be maintained on secure, encrypted servers for a minimum of five years after the publication of our findings in a peer-reviewed journal, and any unanticipated adverse events or risks will be reported by the principal investigator to the Yale Institutional Review Board, which has reviewed and approved this protocol (Protocol Number: 2000035532).

## INTRODUCTION

The current clinical approach for identifying patients with structural heart diseases (SHD) overlooks a critical window where evidence-based medical therapies can be leveraged in asymptomatic stages to improve patient outcomes.^1,2^ The natural history of SHD often includes a long preclinical course before the onset of symptoms, underscoring the critical need for screening strategies to identify patients in this preclinical period.^1,2^ Moreover, large-scale SHD screening using transthoracic echocardiogram (TTE) remains unfeasible due to its unavailability and inaccessibility.^3,4^ Wearable devices are increasingly being recognized as potential tools for enhancing the detection of cardiovascular disease.^5–7^ However, their applicability in clinical practice has yet to be fully understood. In this context, innovative wearable and portable technologies hold promise to improve the management of SHD and patient outcomes through early detection and treatment.

Given the multitude of health-related features, the utility of wearable devices has increased over the last decade, especially for health monitoring.^8–10^ With consistent advancements in healthcare data analytics and biomechanical sensor technology, more sophisticated portable and wearable devices are currently being used for electrocardiogram (ECG) tracing.^11–13^ Furthermore, the use of deep learning on ECGs has been shown to be a promising tool for screening cardiovascular diseases.^14–17^ Therefore, employing deep learning algorithms on ECGs captured by portable devices could significantly broaden the scope for SHD screening at the community level with significant public health implications.^18,19^ Nevertheless, the performance of artificial intelligence-ECG (AI-ECG) algorithms for identifying SHD using portable ECGs for real-world screening is unknown.

We have previously developed and validated a wearable-adapted AI algorithm for detecting SHD, including left ventricular systolic dysfunction (LVSD), valvular disease and severe left ventricular hypertrophy (LVH), from single-lead portable ECGs.^14–16^ This algorithm uniquely maintains its performance despite the common challenge of noise during ECG acquisition with portable devices. In this study, we aim to evaluate the performance of this AI-ECG model in detecting SHD using a wearable and a portable device, each cleared by the US Food & Drug Administration (FDA) for ECG acquisition.^20–23^

### Study Objectives

The primary hypothesis of the study is that our previously developed AI-ECG model can reliably detect SHD using portable ECGs. The primary objective of this study is to assess the performance of the AI-ECG model in detecting SHD using single-lead ECGs obtained from a wearable (Apple Watch) and a portable device among adult patients presenting to the echocardiographic laboratories of Yale New Haven Hospital (YNHH) who undergo an outpatient TTE as part of their routine clinical care.

### Rationale

The use of the AI-ECG model on portable ECGs to detect SHD has not been validated for widespread adoption as a screening tool. Our model was developed exclusively among patients undergoing clinically indicated ECGs as well as TTE within 30 days, using retrospective electronic health records (EHR) data.^14^ This limits the application of this model directly for prospective screening of SHD using portable ECG. Therefore, a thorough evaluation of the model performance for detecting SHD in the healthcare system is needed before clinical adoption.

## RESEARCH METHODS

### Study Design

This is a cross-sectional study with 585 patients referred to the YNHH echocardiographic laboratories for an outpatient TTE as part of their routine clinical care. A poster will be displayed in the echocardiographic laboratory to inform patients about the study **(Appendix 1)**.

The physician in charge will be requested to ask for the willingness of their patients to participate in this study. If the patient expresses interest in participation to their physician, the study coordinator will be notified. Then, the study coordinator will approach the patient and conduct a screening interview to evaluate the patient’s eligibility. The eligible participants will be enrolled in the study after discussing the study objectives and administering written informed consent (**Appendix 2)**.

All study procedures will occur during the hospital visit where the enrolled participants will undergo the previously scheduled routine outpatient TTE. The study coordinator will fill out a short form with the participants information including age, sex, medical record number (MRN) and the use of wearable devices for ECG acquisition in their daily lives in the secure RedCap platform and will obtain a de-identified participant ID. Then, the patient will be asked to use the study portable devices to obtain two separate 30-second ECGs. The study coordinator will add the patient’s unique participant ID to link the ECG signals of the participants to their EHR using MRN-to-Participant ID keys, which will be maintained on Health Insurance Portability and Accountability Act (HIPAA)-compliant platforms. After the study visit, the study coordinator will merge the patient’s clinical data, including demographics, concurrent comorbidities, and the TTE report, from their EHR using their MRN with their portable ECG signals from the company servers using our secure application programming interface (API) and their participant ID.^24^ The study procedures including administration of informed consent and ECGs acquisition will last about five minutes. There will be no follow-up study visits.

### Study Population

The study will involve patients undergoing outpatient TTEs at YNHH as part of their clinical care. Eligible participants for the study must meet the following criteria:

- ≥18 years of age at the time of their TTE.
- Undergo an outpatient TTE as part of routine clinical care.
- Provide signed and dated informed consent.

The following individuals will not be eligible for participation in this study:

- Patients with a cardiac pacemaker or implantable cardioverter defibrillator.
- Pregnant individuals.
- Patients unable to consent due to language barriers, or due to specific impairments that affect communication or cognition.

### Enrollment

Patients who meet all eligibility criteria and sign the informed consent will be enrolled by the study coordinator. In addition to the routine TTE, patients will be instructed to use portable ECG devices for 30 seconds each.

### Study Duration

We expect to complete the study within six months. The study procedures will take approximately five minutes, including protocol discussion, administration of informed consent, and ECG acquisition using the two study devices. All study procedures will occur when the participants present to a YNHH echocardiographic laboratory to undergo a TTE scheduled as a part of their clinical care.

Historically, an average of 791 (±96) unique patients have undergone complete outpatient TTEs at YNHH every month from September 2015 to March 2023 (ranging from 370 in April 2020 to 944 in July 2019). Moreover, the exclusion criteria of the study are minimal (patients with a cardiac pacemaker or implantable cardioverter defibrillator). Additionally, the study involves a non-invasive procedure, TTE, being done as a part of the patient’s routine clinical care, supporting broad participation. Therefore, we believe that the recruitment of 585 individuals who undergo TTE at the YNHH is feasible over six months.

### Study Visit

Participants will have a single study visit, at the time of their TTE. In this study visit:

- The protocol will be explained, and informed consent will be obtained.
- We will ask the participant to use both portable ECG devices as instructed by the study coordinator to record a 30-second ECGs using each device.
- Patients will be monitored for any unanticipated problems during the portable ECG acquisition.

### End of Study and Follow-up

The study will be completed for each participant after the acquisition of the ECGs and there will be no follow-up in-person visits. There will be observational follow up of echocardiographic findings reported by the interpreting cardiologist.

### Randomization

This study does not involve randomization or cohort assignment. All enrolled participants will undergo 2 serial acquisitions of a 1-lead ECG on the two devices following informed consent.

### Outcomes and Analysis

The primary outcome is the performance of the SHD models deployed on 1-lead ECGs captured on wearable and portable devices. This will be defined based on the area under the receiver operating characteristic curve (AUROC), other metrics will include sensitivity, specificity, positive predictive value (PPV), negative predictive value (NPV), accuracy, area under precision-recall curve (AUPRC), and F1-score of the AI-ECG algorithm for detecting SHD using portable ECG devices.

SHD will be defined as the presence of LVSD, valvular disease or severe LVH with TTE being used as the gold standard for confirming these diagnoses. LVSD is characterized by a left ventricular ejection fraction <40% detected on TTE, valvular disease is defined as any moderate or severe stenosis or insufficiency of mitral or aortic valves observed on TTE and severe LVH is defined as a maximal end-diastolic interventricular septal wall thickness of >15 mm with concomitant moderate or severe left ventricular diastolic dysfunction observed on TTE.

### Sample Size and Study Power

We will evaluate the performance of the AI-ECG model for detecting a composite of SHD, that includes LVSD, valvular heart disease and severe LVH. Based on retrospective data from 2015-2022, the overall prevalence of SHD with this definition among patients who undergo an outpatient TTE at the echocardiographic laboratories of YNHH is 15.8%, with a subgroup prevalence of LVSD, valvular disease and LVH of 4.4%, 18.5% and 0.3%, respectively.

The study will be powered to evaluate the composite performance on the broad SHD label (any LVSD, valvular disease or LVH), and explicitly assess the performance on individual disorders of LVSD and valvular disease, which are key for clinical care. Given the extremely low prevalence of severe LVH, the study will not be explicitly powered to detect that subpopulation.

We calculated the sample size based on LVSD, which has a lower prevalence compared with valvular disease, 4.4% versus 18.5%, and will need a higher number of individuals to test the hypothesis with sufficient statistical power. Given (i) 4.4% prevalence of LVSD in the preliminary data, (ii) the AUROC of 0.90 for the AI-ECG model in the published retrospective data,^14^ (iii) a two-sided alpha of 0.05, and (iv) a statistical power of 80%, we will need to recruit 557 individuals to test that the AUROC for detecting LVSD is significantly higher than 0.75 (the lower bound of the confidence interval is higher than 0.75), which is a feasible threshold for a screening model. A sample size of 557 will result in a statistical power of >99.9% to test that the

AUROC is significantly higher than 0.75 for detecting valvular disease with a prevalence of 18.5%.^25,26^ Considering a 5% rate of poor-quality data, we intend to recruit 585 individuals.

### Statistical Design

The study will follow the statistical design principles of a study of a diagnostic test with a known gold standard. For this study we will compare the results of our previously developed wearable-adapted AI algorithm applied to portable ECG signals to the TTE reports, the gold-standard test for diagnosis of SHD.

The primary analysis will calculate the AUROC of the model, the participant level sensitivity or recall, specificity, PPV or precision, NPV, accuracy, F1-score, and AUPRC of the AI-ECG model for detecting SHD and the performance on individual disorders of LVSD and valvular disease. We will report the primary objective variables separately for each portable ECG device, and a composite of them. In the event of missing data necessary for analysis, we will use a multiple imputation approach to minimize bias.

## ETHICS AND DISSEMINATION

### Data Storage and Handling

All data collected by study coordinators from the EHR as a part of initial screening and patient-reported adverse events will be stored on RedCap - Yale’s HIPAA-compliant and secure data collection platform with a built-in audit trail or other HIPAA-compliant servers at Yale. The database servers are only accessible from within the Yale intranet (or via virtual private network [VPN] remotely) and additionally requires a separate login username and password and multifactor authentication. No protected health information will be included in the analysis or publications. All analyses will proceed on these servers alone. The linking file will be maintained for potential future linking to national databases.

Data will be maintained on secure, encrypted servers for a minimum of five years after the publication of our findings in a peer-reviewed journal (in such case as there is a need to return to the original source data to validate a finding or respond to a question).

### Human Subject Protection

The principal investigator is responsible for monitoring the data, assuring protocol compliance, and conducting safety reviews at a weekly frequency. During the review process, the principal investigator will evaluate whether the study should continue unchanged, require modification/amendment, or stop enrollment. The principal investigator and the IRB have the authority to stop or suspend the study or require modifications.

We will use two FDA-cleared portable ECG devices, that were not associated with any adverse event either in clinical studies or in post-marketing reports. Therefore, we do not anticipate adverse outcomes justifying study discontinuation. However, the principal investigator and the IRB have the right to discontinue the study in case of unanticipated problems that occur during the study.

### Informed Consent

All participants will be required to provide signed informed consent before participation. Informed consent will be taken before the enrollment of participants for using the portable ECG devices. The proposed informed consent form is included in **Appendix 2**.

Study coordinators will facilitate informed consent procedures, allowing participants sufficient uninterrupted time for review and clarification. To confirm comprehension, participants will be encouraged to explain the procedure and its risks in their own words. Open-ended questions will be used to ensure a thorough understanding of the study protocol.

### Subject Confidentiality

Subject confidentiality is held in strict trust by the research team. Subject medical record review will be limited to the elements needed to complete the study. Only authorized HIPAA- and Good Clinical Practice (GCP)-trained study team members can extract research data from medical records. Age, sex, and key health information measures (provider encounters, notes, comorbidities, medication lists, problem lists, family history, allergies, laboratory findings, procedures, immunizations, vital signs, imaging and other diagnostic testing, and medical record numbers) will be collected from the EHR and de-identified before analysis.

### Participant Compensation

We will conduct a lottery to reward ten participants with a prepaid Visa gift card worth $25 each.

### Management and Reporting of Adverse Events

Despite the minimal risk nature of the study, participants will be asked to report any experienced adverse events to the study coordinator and this information will be recorded. The investigators will determine whether the adverse event was likely to be related to the study or not. For events deemed to be study-related, the investigators will categorize them as serious or not depending on whether the event required a visit to a healthcare provider (office visit, urgent care, emergency care, or other) or the use of prescription medication. Serious related adverse events will be reported to the IRB within three business days of investigators becoming aware of the event. Unrelated and non-serious, related adverse events will be recorded and reported to the companies providing the study devices.

### Institutional Review Board Approval

The protocol was reviewed and approved (Protocol Number: 2000035532) by the Yale IRB. Any changes to the protocol or study team will require approval from the IRB before implementation. The IRB will conduct a continuing review at intervals appropriate to the degree of risk, but not less than once per year. A study closure report will be submitted to the IRB after all research activities have been completed. Other study events (e.g., data breaches, protocol deviations) will be submitted per Yale’s IRB’s policies.

### Plans for Disseminating Study Results

The study will be conducted in accordance with the ethical principles that have their origin in the Declaration of Helsinki, Common Rule, GCP, IRB, and applicable record retention policies. All data are accessed through YNHH EHR which is HIPAA compliant. A manuscript describing the primary objective once the research study has been completed will be submitted for publication and/or presented at a scientific meeting. Authorship will be determined by mutual agreement and in line with the International Committee of Medical Journal Editors authorship requirements.

### Patient and Public Involvement

Patients or the general public were not involved in the design of the study protocol.

## Data Availability

All data produced in the present study are available upon reasonable request to the authors.

## APPENDIX 1 Join our study to advance cutting-edge medical technology in just 5 minutes!

Help us study a groundbreaking technology for detecting structural heart disease using portable smart devices recording your electrocardiogram!

If you are interested, please ask your physician at the Echocardiogram Laboratory and join the lottery!

In this study, we are looking for volunteers undergoing echocardiogram as part of their routine clinical care at th e Echocardiogram Laboratory of Yale-New Haven Hospital who are willing to wear two portable smart devices for 30 seconds to record their electrocardiogram.

## APPENDIX 2 COMPOUND AUTHORIZATION AND CONSENT FOR PARTICIPATION IN A RESEARCH STUDY

**YALE UNIVERSITY SCHOOL OF MEDICINE**

**Study Title:**

**Performance of a Wearable-Adapted Artificial Intelligence-Driven Algorithm for Detecting Cardiomyopathies Using Portable Electrocardiogram**

**Principal Investigator (the person who is responsible for this research):**

Rohan Khera, MD, MS, 195 Church Street, 6^th^ Floor New Haven, CT 06510, Telephone Number: (319) 400-6261 Email Address

### Research Study Summary

- You are being consented to join a research study.
- The purpose of this study is to assess the performance of an artificial intelligence (AI) algorithm to detect cardiomyopathies, a type of structural heart disease, using the electrocardiogram (ECG) obtained by portable ECG devices.
- For this study, in addition to your routine clinical care, including but not limited to an echocardiogram, you will be asked to use one wearable device (Apple Watch) and one 1-lead portable ECG device for 30 seconds.
- There is minimal risk associated with participating in this study. The AI algorithm will be applied to the ECG signals obtained by portable ECG devices. Hence, no risk is posed to the participant with the evaluation of their ECG by the AI algorithm. The portable ECG devices that will be used in this study have been cleared by the US Food and Drug Administration (FDA) for ECG acquisition.
- There are no expected immediate benefits to the participants of this study; however, societal benefits from the development of an accurate screening tool for cardiomyopathies may be significant.
- Taking part in this study is your choice. You can choose to take part, or you can choose not to take part in this study. You can also change your mind at any time. Whatever choice you make, you will not lose access to your medical care or give up any legal rights or benefits.
- If you are interested in learning more about the study, please continue reading, or have someone read to you, the rest of this document. Take as much time as you need before you make your decision. Ask the study staff questions about anything you do not understand. Once you understand the study, we will ask you if you wish to participate. If so, you will have to sign this form.

**Why is this study being offered to me?**

You are being requested to participate in the study as you are going to receive an echocardiogram as part of your routine clinical care at the Echocardiographic Laboratory of Yale-New Haven Hospital. We have taken approval from your care provider for requesting your participation in our study. If you choose to participate in the study, we will obtain your ECG using two portable ECG devices to assess the performance of a novel AI algorithm in detecting cardiomyopathies, a type of structural heart disease, using your portable ECG. We are looking for 585 participants to be part of this study.

**Who is paying for the study?**

This study is funded by the Doris Duke Charitable Foundation, awarded to the principal investigator, Dr. Rohan Khera, MD, MS.

**What is the study about?**

The purpose of this study is to assess the performance of a novel AI algorithm, to detect cardiomyopathies using the ECG obtained by portable ECG devices.

**What are you asking me to do and how long will it take?**

If you agree to take part in this study, this is what will happen:

In addition to your routine clinical care, you will be asked to use two FDA-cleared portable ECG devices, for 30 seconds each.

**What are the risks and discomforts of participating?**

There is minimal risk from participating in this study. The AI algorithm will be applied to the ECG signals obtained by portable ECG devices. Therefore, it does not pose any risk to the participant. Moreover, no adverse events were observed during the clinical testing of the devices used in the study. These devices have been cleared by the FDA for ECG acquisition.

The minimum time for the acquisition of an interpretable ECG signal is 30 seconds, and you will use these devices for this minimal time after giving consent.

Individuals with pacemakers and/or implantable cardioverter-defibrillators will not be included to minimize the risk since the portable device used in this study has not been tested or recommended for use in these individuals.

**How will I know about new risks or important information about the study?**

The study will last around five minutes other than obtaining the informed consent and will end with your current health encounter. However, we will contact you through your healthcare provider at the Echocardiogram Laboratory if we learn any new information that could be of interest to you.

**How can the study possibly benefit me?**

There are no expected immediate benefits to the participants of this study; nevertheless, the possible implication of using smart devices for detecting cardiomyopathies, will have important clinical and public health implications. The primary benefit of this study will be societal.

**How can the study possibly benefit other people?**

The benefits to science and other people may include the application of the novel AI algorithm to improve cardiovascular care and health outcomes. The results of this study can benefit the society-at-large as the AI algorithm may be used as a screening tool for cardiomyopathies in the future. In this case, many individuals can take advantage of this algorithm for timely diagnosis and treatment for these conditions, which improves their quality of life and health outcomes.

**Are there any costs to participation?**

If you take part in this study, you will not have to pay for any services, supplies, or study procedures.

**Will I be paid for participation?**

We will be awarding 10 participants in the lottery with a $25 prepaid Visa gift card each.

**What are my choices if I decide not to take part in this study?**

You can choose to not participate in the study. It will not affect your routine clinical care.

**How will you keep my data safe and private?**

We will keep the information we collect about you confidential. We will share it with others if you agree to it or when we have to do it because U.S. or State law requires it. For example, we will tell somebody if we learn that you are hurting a child or an older person. Your data will be stored in a secure, password-protected server with a study-specific ID number. Only study personnel and regulators would have access to this data. When we publish the results of the research or talk about it at conferences, we will not use your name. If we want to use your name, we will ask you for your permission. We will also share information about you with other researchers for future research, but we will not use your name or other identifiers. We will not ask you for any additional permission. Your data may be used for future research studies or distributed to another investigator for future research studies without additional informed consent from you. In the event that such data is shared for future studies, all identifiable information will be removed.

**What information will you collect about me in this study?**

The information we are asking to use and share is called “Protected Health Information.” It is protected by a federal law called the Privacy Rule of the Health Insurance Portability and Accountability Act (HIPAA). In general, we cannot use or share your health information for research without your permission. If you want, we can give you more information about the Privacy Rule. Also, if you have any questions about the Privacy Rule and your rights, you can speak to Yale Privacy Officer at 203-432-5919.

The specific information about you and your health that we will collect, use, and share include:

- Research study records
- Medical and laboratory records of ECG were done in connection with this study.
- Records about phone calls made as part of this research
- Information obtained during this research regarding

■ Echocardiogram reports
■ Results from the medical chart, including past medical history, laboratory, x-ray, and other test results
■ Physical exam reports
- The entire research record

We will retrieve your medical records up to the index health encounter in which you undergo an echocardiogram.

**How will you use and share my information?**

We will use your information to conduct the study described in this consent form. We may share your information with:

- The U.S. Department of Health and Human Services (DHHS) agencies
- The FDA, so that the FDA can review information about the algorithm. The information may also be used to meet the reporting requirements of drug regulatory agencies.
- Representatives from Yale University, the Yale Human Research Protection Program, and the Institutional Review Board (the committee that reviews, approves, and monitors the research on human participants), who are responsible for ensuring research compliance. These individuals are required to keep all information confidential.
- Health care providers who provide services to you in connection with this study.
- Laboratories and other individuals and organizations that analyze your health information in connection with this study, according to the study plan.
- Principal investigator of the study
- Co-Investigators and other investigators
- Study coordinator and members of the research team

We will ensure your information stays private. But, if we share information with people who do not have to follow the Privacy Rule, your information will no longer be protected by the Privacy Rule. Let us know if you have questions about this. However, to better protect your health information, agreements are in place with these individuals and/or companies that require that they keep your information confidential.

**Why must I agree to the information in this document?**

By agreeing, you will allow researchers to use and disclose the information described above for this research study. This is to ensure that the information related to this research is available to all parties who may need it for research purposes. You always have the right to review and copy your health information in your medical record.

**What if I change my mind?**

The authorization to use and disclose your health information collected during your participation in this study will never expire. However, you may withdraw or take away your permission at any time. You may withdraw your permission by telling the study staff or by writing to Rohan Khera, MD, MS, 195 Church Street, 6^th^ Floor, New Haven, CT 06510.

If you withdraw your permission, you will not be able to stay in this study, but the care you get from your doctor outside this study will not change. No new health information identifying you will be gathered after the date you withdraw. Information that has already been collected may still be used and given to others until the end of the research study to ensure the integrity of the study and/or study oversight.

**Who will pay for treatment if I am injured or become ill due to participation in the study?**

If you are injured while on study, seek treatment and contact the study doctor as soon as you are able.

Yale School of Medicine and Yale-New Haven Hospital do not provide funds for the treatment of research-related injury. If you are injured as a result of your participation in this study, treatment will be provided. You or your insurance carrier will be expected to pay the costs of this treatment. No additional financial compensation for injury or lost wages is available. You do not give up any of your legal rights by signing this form.

**What if I want to refuse or end participation before the study is over?**

Taking part in this study is your choice. You can choose to take part, or you can choose not to take part in this study. You also can change your mind at any time. Whatever choice you make, you will not lose access to your medical care or give up any legal rights or benefits. Not participating or withdrawing later will not harm your relationship with your own doctors or with this institution.

To withdraw from the study, you can ask the study coordinator at any time and tell them that you no longer want to take part.

**What will happen with my data if I stop participating?**

If you stop participating, your data will be fully de-identified. Data collected prior to the time you stop participating would be retained in de-identified form.

**Who should I contact if I have questions?**

Please feel free to ask about anything you don’t understand.

If you have questions later or if you have a research-related problem, you can call the Study Coordinator at (203) 747-4429.

If you have questions about your rights as a research participant, or you have complaints about this research, you call the Yale Institutional Review Board at (203) 785-4688 or.

### Authorization and Permission

Your signature below indicates that you have read this consent document and that you agree to be in this study. We will give you a copy of this form.

|  |  |  |
| --- | --- | --- |
| Participant Printed Name | Participant Signature | Date |
| Person Obtaining Consent Printed Name | Person Obtaining Consent Signature | Date |

